# Reconstructing the early global dynamics of under-ascertained COVID-19 cases and infections

**DOI:** 10.1101/2020.07.07.20148460

**Authors:** Timothy W. Russell, Nick Golding, Joel Hellewell, Sam Abbott, Lawrence Wright, Carl A B Pearson, Kevin van Zandvoort, Christopher I Jarvis, Hamish Gibbs, Yang Liu, Rosalind M. Eggo, W. John Edmunds, Adam J. Kucharski, on behalf of the CMMID COVID-19 working group

**Affiliations:** Centre for Mathematical Modelling of Infectious Diseases, London School of Hygiene & Tropical Medicine; Telethon Kids Institute and Curtin University, Perth, Western Australia; Defence Science and Technology Laboratory/Sopra Steria, Fareham, United Kingdom

**Keywords:** Case ascertainment, COVID-19, SARS-CoV-2, surveillance, under-reporting, situational awareness, outbreak analysis

## Abstract

**Background:** Asymptomatic or subclinical SARS-CoV-2 infections are often unreported, which means that confirmed case counts may not accurately reflect underlying epidemic dynamics. Understanding the level of ascertainment (the ratio of confirmed symptomatic cases to the true number of symptomatic individuals) and undetected epidemic progression is crucial to informing COVID-19 response planning, including the introduction and relaxation of control measures. Estimating case ascertainment over time allows for accurate estimates of specific outcomes such as seroprevalence, which is essential for planning control measures.

**Methods:** Using reported data on COVID-19 cases and fatalities globally, we estimated the proportion of symptomatic cases (i.e. any person with any of fever >= 37.5°C, cough, shortness of breath, sudden onset of anosmia, ageusia or dysgeusia illness) that were reported in 210 countries and territories, given those countries had experienced more than ten deaths. We used published estimates of the baseline case fatality ratio (CFR), which was adjusted for delays and under-ascertainment, then calculated the ratio of this baseline CFR to an estimated local delay-adjusted CFR to estimate the level of under-ascertainment in a particular location. We then fit a Bayesian Gaussian process model to estimate the temporal pattern of under-ascertainment.

**Results:** Based on reported cases and deaths, we estimated that, during March 2020, the median percentage of symptomatic cases detected across the 84 countries which experienced more than ten deaths ranged from 2.4% (Bangladesh) to 100% (Chile). Across the ten countries with the highest number of total confirmed cases as of 6th July 2020, we estimated that the peak number of symptomatic cases ranged from 1.4 times (Chile) to 18 times (France) larger than reported. Comparing our model with national and regional seroprevalence data where available, we find that our estimates are consistent with observed values. Finally, we estimated seroprevalence for each country. As of the 7th June, our seroprevalence estimates range from 0% (many countries) to 13% (95% CrI: 5.6% – 24%) (Belgium).

**Conclusions:** We found substantial under-ascertainment of symptomatic cases, particularly at the peak of the first wave of the SARS-CoV-2 pandemic, in many countries. Reported case counts will therefore likely underestimate the rate of outbreak growth initially and underestimate the decline in the later stages of an epidemic. Although there was considerable under-reporting in many locations, our estimates were consistent with emerging serological data, suggesting that the proportion of each country’s population infected with SARS-CoV-2 worldwide is generally low.

**Funding:** Wellcome Trust, Bill & Melinda Gates Foundation, DFID, NIHR, GCRF, ARC.

## Background

The pandemic of the novel coronavirus SARS-CoV-2 has caused 25.3 million confirmed cases and 846,841 deaths as of 31^h^ August 2020 (1). As a precautionary measure, or in response to locally detected outbreaks, countries have introduced control measures with varying degrees of stringency (1), including isolation and quarantine; school and workplace closures; bans on social gatherings; physical distancing and face coverings; and stay-at-home orders (2,3). Several features of SARS-CoV-2 make accurate detection during an ongoing epidemic challenging (4-6), including high transmissibility (3,7,8); an incubation period with a long-tailed distribution (9); pre-symptomatic transmission (10); and the existence of asymptomatic infections, which may also contribute to transmission (11). These attributes mean that infections can go undetected (12) and that countries may only detect and report a fraction of their infections (3,13).

Understanding the extent of unreported infections in a given country is crucial for situational awareness. If the true size of the epidemic can be estimated, this enables a more reliable assessment of how and when non-pharmaceutical interventions (NPIs) should be both introduced, as infections rise, or relaxed as infections fall (3). Estimates of infection prevalence are also important for obtaining accurate measures of transmission: if the proportion of infections reported declines as the epidemic rises, the number of confirmed cases will grow slower than the actual underlying epidemic. Likewise, if detection rises as the epidemic declines, it may appear that transmission is not declining as fast as it is in reality. Underdetection of cases also makes it challenging to estimate at what stage of the epidemic a particular country is (14): viewed in isolation, case incidence data could reflect a very large undetected epidemic, or a smaller, better reported epidemic.

To estimate how the levels of under-ascertainment vary over time, we present a modelling framework that combines data on reported cases and deaths, and published severity estimates. We apply our methods to countries that have reported more than ten deaths to date, then use these under-ascertainment estimates to reconstruct global epidemics in all countries where case and death time series data are available. We also compare the model estimates for cumulative incidence against existing seroprevalence results. Finally, we present the adjusted case curves for the ten countries with the highest confirmed and adjusted case numbers, as well as global prevalence estimates for SARS-CoV-2.

## Methods

As SARS-CoV-2 infections that generate mild symptoms are more likely to be missed than severe cases, the ratio of cases to deaths, adjusting for delays from report to fatal outcome, can provide information on the possible extent of undetected symptomatic infections. Using a Bayesian Gaussian process model, we estimate changes in under-ascertainment over time, as described below.

### Adjusting for delay from confirmation to death

In real time, simply dividing deaths to date by cases to date leads to a biased estimate of the case fatality ratio (CFR), because this naive calculation does not account for delays from confirmation of a case to death, and under-ascertainment of cases (5,6) and in some circumstances, under-ascertainment of deaths too. Using the distribution of the delay from hospitalisation to death for cases that are fatal, we can estimate how many cases so far are expected to have known outcomes (i.e. death or recovery), and hence adjust the naive estimates of CFR to account for these delays and produce a delay-adjusted CFR (dCFR). Separately published dCFR estimates for a given country can be used to estimate the number of symptomatic cases that would be expected for a given dCFR trajectory. Available estimates for the CFR that adjust for under-reporting typically range from 1-1.7% (7-10). Large studies in China and South Korea estimate the CFR at 1.38% (95% CrI: 1.23-1.53%) (9) and 1.7% (95% CrI: 1.1-2.5%) (7) respectively.

### Inferring level of under-ascertainment

Assuming a baseline CFR of 1.4% (95% CrI: 1.2% – 1.5%), the ratio of this baseline CFR to our estimate of the dCFR for a given country can be used to derive a crude estimate of the proportion of symptomatic cases that go unreported for this country. For each country we calculate the dCFR on each day and use the ratio of the baseline CFR to the dCFR estimate to produce daily estimates of the proportion of unreported cases. We then use a Gaussian process (GP) model to fit a time-dependent under-ascertainment rate for each country. A more detailed description of the methods, including the mathematical details of the Gaussian process and the different sources of uncertainty present in the model, can be found in the Supplementary Material.

With the aim of developing a parsimonious and easily transferable analysis framework, we assume the same baseline CFR for all countries in the main results. Given that CFR varies substantially with age (5), this induces a certain amount of error in our estimates, especially for countries with age distributions significantly different to China, where the data used to derive the baseline CFR estimates originated (5). Therefore, we include a version of all the main results where we compute an indirectly adjusted baseline CFR, using the underlying age distribution of each country using the wpp2019 R package (15) and the age-stratified CFR estimates from (5) in the supplementary material (Additional file 1: Figures S5, S6 and S7), where we also include a verbose limitations section discussing at length the potential errors induced under such assumptions.

### Relationship between under-ascertainment and testing

We attempt to characterise the relationship between widespread RT-PCR testing and case ascertainment using our temporal under-ascertainment estimates and testing data for many countries from OurWorldInData (16). We do so by performing a correlation test between the two for all countries we had both data for. The resulting bivariate scatterplot is included in the supplementary material (Additional file 1: Figure S3).

### Comparison against seroprevalence estimates

We attempted to reconstruct the infection curves by first adjusting the reported case data for under-ascertainment (Figure 1). We then adjust further these estimated symptomatic case curves so that they represent all infections. We do so using the assumption that 50% of infections are asymptomatic overall, with an assumed wide range between 10% – 70% and mean-lagging the time point to adjust for the delay between onset of symptoms to confirmation (17). We assume that serological tests are broadly similar between locations, in order for a tractable and relatively simple comparison. We include both our estimates, with their 95% credible intervals, and the confidence intervals of all serological estimates in our comparison (Figure 3).

**Figure 1:**
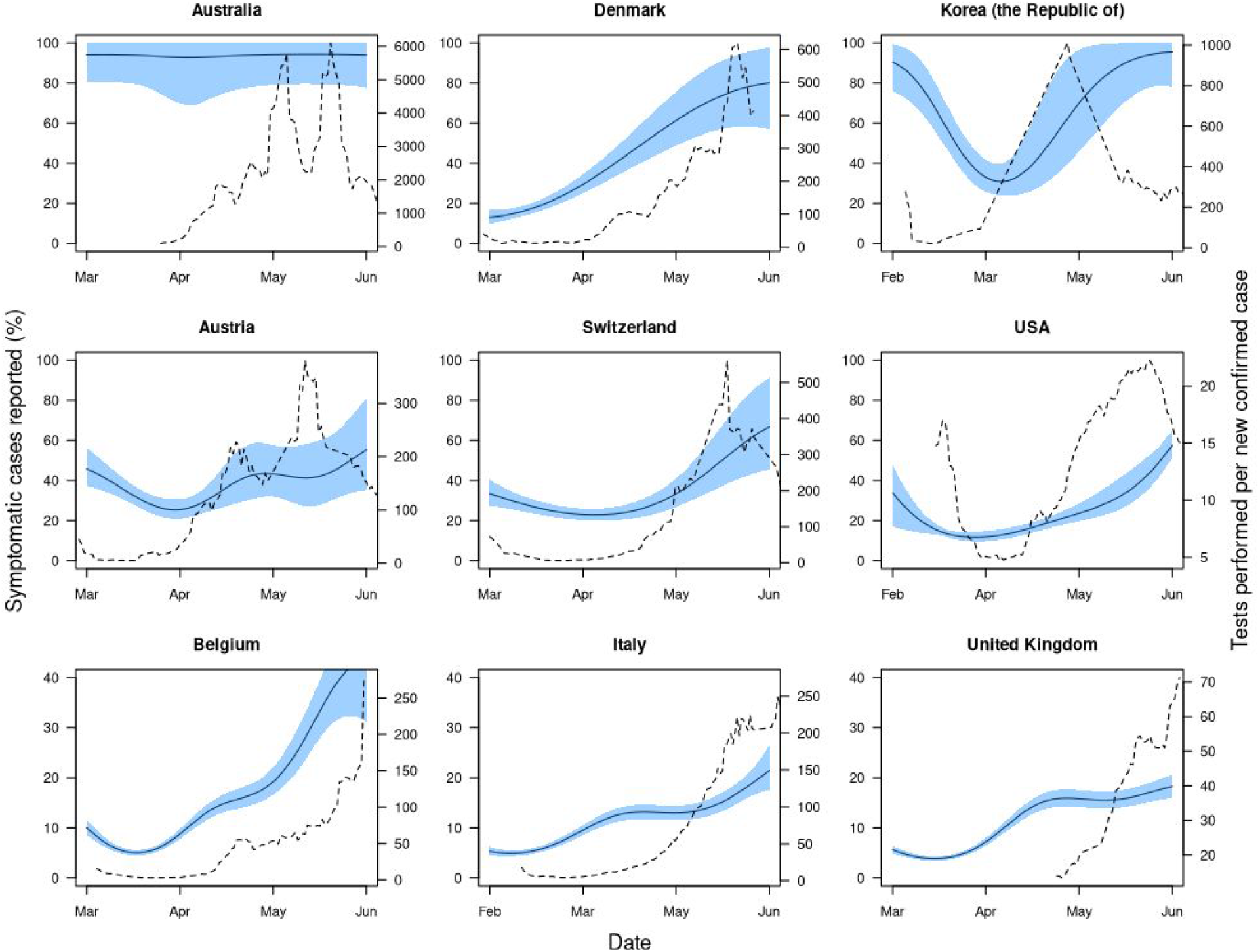
Illustrative examples of temporal variation in under-ascertainment and testing effort. Nine countries under-ascertainment and testing effort dynamics, where the under-ascertainment dynamics display a typical U-trend. The solid black line is the estimated median proportion of symptomatic cases ascertained over time and the shaded blue region is the 95% credible interval of these ascertainment estimates. Dashed line shows the reported testing effort, which we defined as a 7-day moving average of the number of new tests per new case reported each day. The illustrative examples chosen in Figure 1 were constrained by the availability of testing data over a time period comparable to our under-ascertainment estimates. However, all countries under-ascertainment estimates, with or without testing data, are presented in Additional file 1: Figure S1.

### Data and code availability

The data we use is publicly available online from the European Centre for Disease Control (ECDC) (18). The code for the dCFR and under-reporting estimation model can be found here: https://github.com/thimotei/CFR_calculation. The code to read in the under-ascertainment data and to reproduce the figures in this analysis can be found here: https://github.com/thimotei/covid_underreporting.

## Results

We estimated substantial variation in the proportion of symptomatic cases detected over time in many of the countries considered (Figure 1 & Figure S1). For example, during March the median percentage of symptomatic cases detected across the 84 countries which experienced more than ten deaths ranged from 2.38% (Bangladesh) to 99.6% (Chile). Also during March, the median percentage of symptomatic cases detected across Europe ranged from 4.81% (France) to 85.5% (Cyprus).

Countries might expect to detect an increasing proportion of symptomatic cases if they scale up testing effort in response to the outbreak. To measure this, we compared our estimates for the proportion of cases detected with the number of tests performed per new case each day, which can provide an indication of testing effort with a country (18). Taking a moving average with a 7-day window, we found that countries that showed high testing effort did not necessarily have high levels of case ascertainment. For example, in a two-week period in March the United Kingdom performed 80 tests per new case (the mean across Europe during the same period was 27 tests per new case). However, we estimate that also in the UK only between 3-10% of symptomatic cases were being detected (Figure 1). Overall, we found a weak positive correlation between testing effort and case ascertainment (Kendall’s correlation coefficient of 0.16). This suggests that increased testing effort can help to improve case ascertainment, but on its own is not enough to guarantee low levels of under-ascertainment.

Using our temporal under-ascertainment trends, we estimate that during March, April, and May the percentage of symptomatic cases detected in European countries and averaged over time ranged from 4.8% – 86% (France – Cyprus), 5.8% – 100% (France – Belarus) and 11% – 86% (Hungary – Cyprus) respectively. By comparison, the number of reported tests performed per new confirmed case, averaged over the month in question, ranged between 2.7 to 76 in March (Belgium – Portugal), 2.7 to 832 in April (Belgium – Slovakia) and 12 to 1334 (Ukraine – Lithuania) in May.

Adjusting confirmed case data for under-ascertainment to obtain estimated symptomatic case curves, we found a much larger and more peaked epidemic in the ten countries with the highest total number of confirmed cases and the ten with the highest number of adjusted cases as of 6th July 2020 (Figure 2, with estimates for other countries shown in Additional file 1: Figure S2). Typically, the estimated peak of symptomatic cases in these countries ranged from 1.4 times (Chile) to 17.8 times larger (France) than the peak in the reported case data (Table 1). Moreover, in the five countries of these ten that had a clear initial peak before the end of May 2020, we estimated that the post-peak decline in the number of infections was steeper than that implied by the confirmed case curves (Figure 2B).

**Table 1:** Comparison between the confirmed and adjusted case numbers at their respective peaks for ten countries with the highest number of total confirmed cases and ten countries with the highest number of symptomatic cases after adjusting for under-ascertainment. Eight countries are in both lists, so the total is twelve distinct countries. We find that the peak of the case curves shifts when they are adjusted for under-ascertainment. Clearly, Mexico and Brazil haven’t necessarily peaked yet, given that they are not as far along their epidemic as the other countries. Therefore, for these countries, we simply report the date and number of the highest number of cases to-date.

|  | Date |  | Value at peak |  |
| --- | --- | --- | --- | --- |
| Location | Peak of confirmed cases | Estimated change in peak date (absolute value) | New confirmed cases at peak | Estimated total cases (95% CrI) |
| Brazil | 6th June 2020 | 0 days | 54,771 | 122,512 (110,660 - 137,374) |
| Chile | 18th June 2020 | 3 days | 36,179 | 52,042 (47,828 - 56,338) |
| France | 1st April 2020 | 0 days | 7,578 | 134,594 (120,450 - 151,352) |
| India | 21st June 2020 | 18 days | 15,413 | 48,513 (43,433 - 54,939) |
| Iran | 5th April 2020 | 0 days | 5,275 | 17,931 (16,078 - 20,201) |
| Italy | 22nd March 2020 | 0 days | 6,557 | 75,521 (64,229 - 91,630) |
| Mexico | 13th June 2020 | 0 days | 5,222 | 55,661 (50,204 - 62,237) |
| Peru | 4th June 2020 | 0 days | 24,603 | 24,603 (22,121 - 27,629) |
| Russia | 12th June 2020 | 4 days | 11,656 | 15,604 (14,248 - 17,270) |
| Spain | 27th March 2020 | 1 day | 9,181 | 85,881 (77,697- 96,319) |
| UK | 12th April 2020 | 0 days | 8,719 | 100,870 (91,054 - 112,639) |
| USA | 26th April 2020 | 21 days | 48,529 | 280,631 (226,097 - 344,472) |

**Figure 2:**
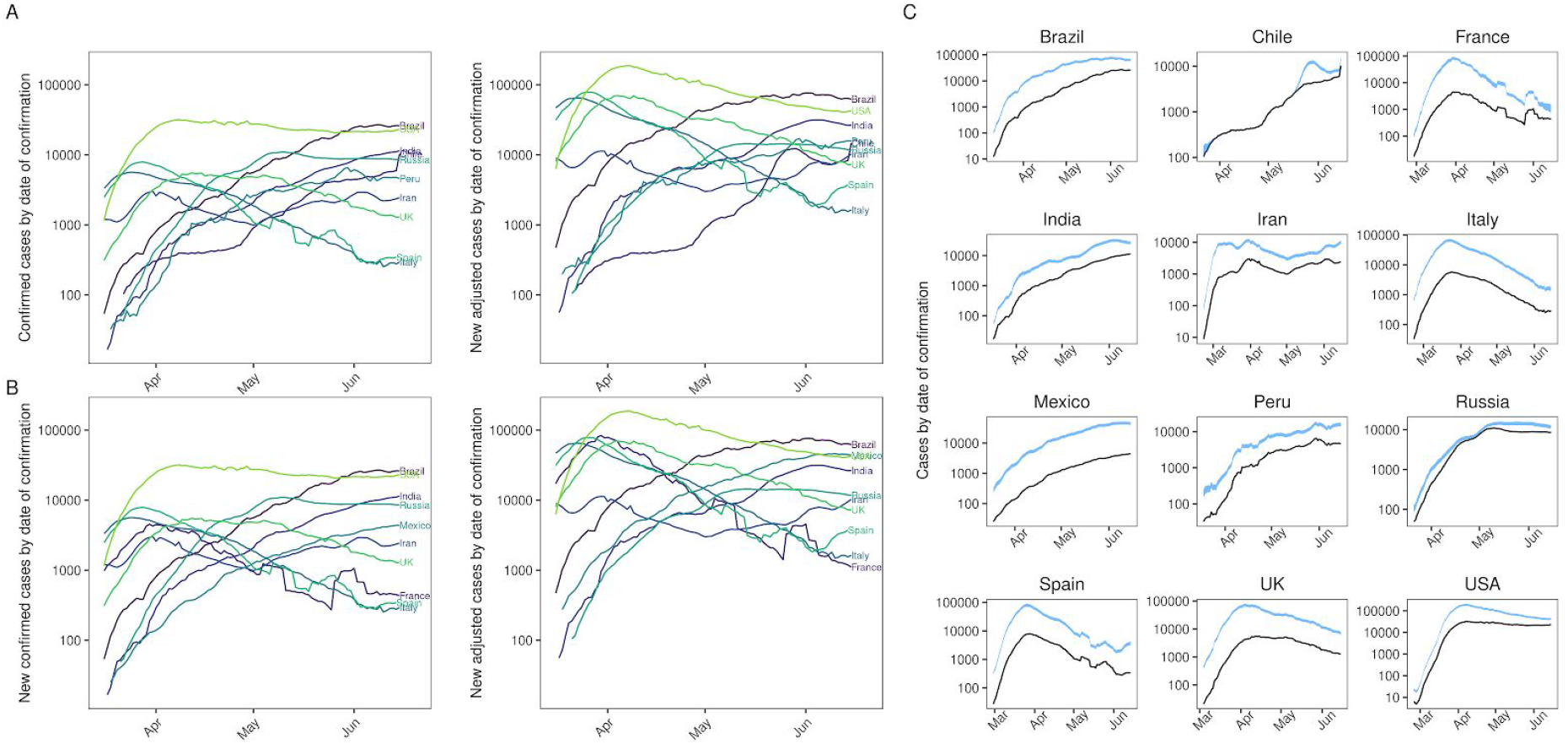
Confirmed case curves adjusted for temporal under-ascertainment. Panel A: Confirmed cases (left) and adjusted cases (right) for the ten countries with the highest number of confirmed cases. Panel B: Confirmed cases (left) and adjusted cases (right) for the ten countries with the highest number of confirmed cases after adjusting for under-ascertainment. There are two countries which change between panels A and B: France and Mexico are replaced by Chile and Peru respectively. Panel C: The same curves plotted in panel A, but with a plot per country. Blue shaded region corresponds to the 95% CrI of the adjusted curves. Panels A and B highlight between country variation whereas panel C highlights within country variation.

We also compared the estimated proportion of individuals infected in our model with seroprevalence studies that measured the prevalence of SARS-CoV-2 antibodies. We represent our cumulative incidence estimates in the same form as the observed serological estimates, as a percentage of the population. This is either the population of the country or the population of some smaller region or sub-region, depending on the serological dataset. We found that all but one of the published seroprevalence estimates fell within the 95% credible interval (CrI) of our estimated cumulative incidence curves over time, with the one exception being Denmark where we underestimated the observed seroprevalence (Figure 3).

**Figure 3:**
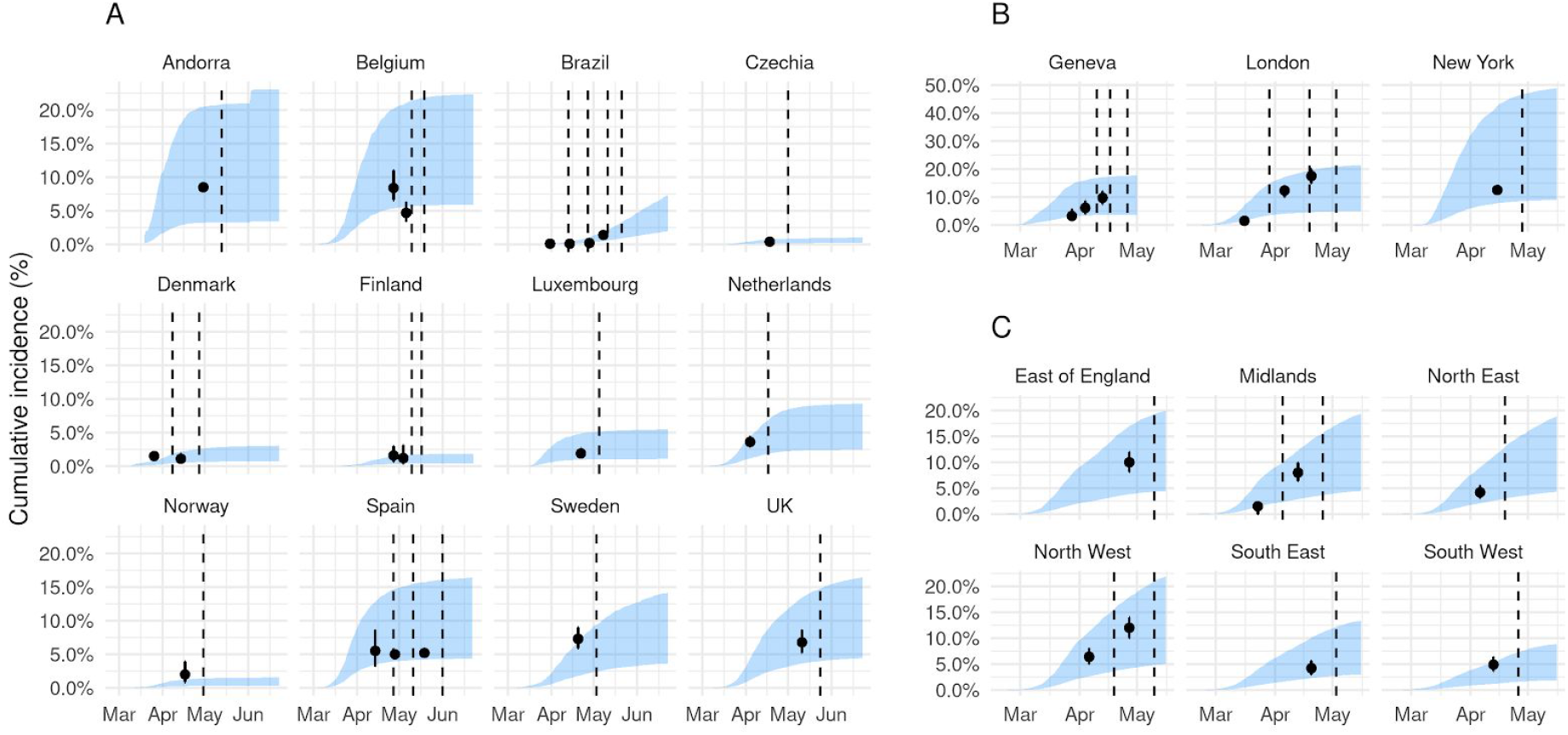
Estimated infection prevalence curves compared with observed seroprevalence data. Panel A: country-level comparisons. Panel B: City-level comparisons for Geneva, London and New York. Panel C: Regional-level comparisons, using six of the eight regions of England. North West and Yorkshire are aggregated together and London is shown above in Panel B: After adjusting the reconstructed new cases per day curves for potential asymptomatic infections and for the delay between onset of symptoms and confirmation, we sum up the cases and divide by the population in each country or region, to estimate the total percentage infected. We are then able to directly compare the model estimates to existing seroprevalence results (black points, with 95% binomial CI above and below). Dashed line shows the end of the serological testing period, therefore we lag the seroprevalence estimate by the mean delay between infection-to-seroconversion, which is likely to be around 14 days (19).

Applying our estimation method to all countries for which case and death time series data are available, we produced a map of seroprevalence estimates as of 16th June (Figure 4A), suggesting that most infections by this point had been concentrated in Europe and the US. We estimate that between 0.02% – 15% of populations in Europe have been infected. As of the middle of May, cases were in Latin America and Africa. For both continents combined, we estimate that between 0.00% – 3.48% of the population of these two continents had been infected as of 16 June 2020. We also reconstructed the early progression of the COVID-19 pandemic across Europe (Figure 4B), finding that the estimated infection prevalence over time was an order of magnitude higher than the confirmed case numbers suggest, with prevalence growing rapidly in late February and early March in several countries.

**Figure 4:**
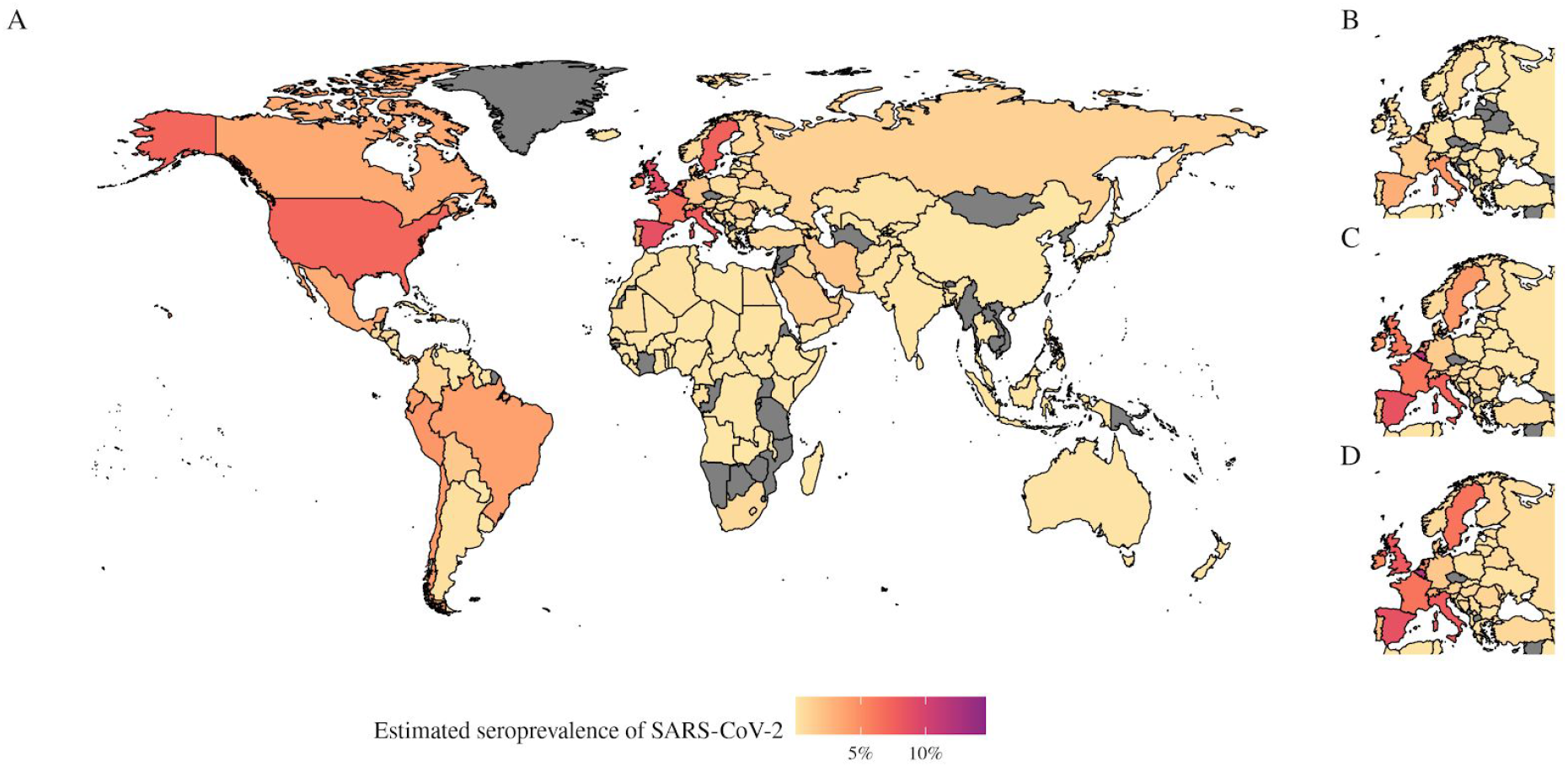
Map of estimated seroprevalence as of the start of June, where we adjusted for under-ascertainment of symptomatic cases and asymptomatic infections. A) Estimated seroprevalence of SARS-CoV-2 globally as of 7th June 2020, for all countries we have reliable estimates for – greyed out countries represent where we did not have reliable estimates due to insufficient data. B-D) The estimated seroprevalence of SARS-Cov-2 in Europe on B) 31st March, C) 30th April and D) 31st May. represent where there was insufficient data to compute reliable estimates.

## Discussion

The epidemiological and clinical characteristics of SARS-CoV-2 mean that a large proportion of infections may go undetected (13,20). In the absence of serological data, the ratio between cases and deaths, adjusted for delays from confirmation-to-outcome, can be used to derive estimates of the proportion of symptomatic cases reported. Using this approach, we estimated that case ascertainment dropped substantially in many countries during the peak of their first epidemic wave. Although serological surveys are beginning to emerge (20), many countries do not have such data available, or may only have results from a single cross-sectional survey. The methods and estimates presented here can therefore provide an ongoing picture of the underlying epidemics, including local level dynamics as fine-scale surveillance data become available (21,22).

Our analysis has some limitations. We assumed the age-adjusted baseline CFR was 1.4% (95% CrI: 1.2% – 1.5%) (4), which is broadly consistent with other published estimates (5,23,24), and we assumed a range of 10% – 70% of infections were asymptomatic (20,25,26) with a mean value of 50% (12). Given the uncertainty in these estimates, we propagated the variance in baseline CFR and range in proportion asymptomatic in the inference process so the final 95% credible interval reported for under-ascertainment reflects underlying uncertainty in the model parameters. We also assumed that deaths from COVID-19 are accurately reported. If local testing capacity is limited, or if testing policy affects attribution of deaths (for example, the evidence for the efficacy of post-mortem swabbing is lacking), deaths can be misattributed to a cause other than COVID-19. In that case, our model may underestimate the true burden of infection. For example, in Peru between 1st April and 1st July 2020, there were excess deaths when compared to confirmed COVID deaths and 3396 reported COVID-19 deaths per 100,000 cases, whereas in the United Kingdom there were 199% excess deaths, and 23,642 reported COVID-19 deaths per 100,000. There have also been reports of data reporting issues for several countries (27). Additionally, if a large proportion of transmission is concentrated within specific age groups, the effective CFR may be higher or lower than the assumed baseline; with better age-stratified temporal data on cases and deaths, it would be possible to explore the effect of this in more detail. However, our estimates were in general consistent with published serological data, where available, providing evidence that our method was robust for these countries at least.

To compare our estimates against seroprevalence studies, and consistent with other simplifying assumptions across countries in this study, we assume that there is little or no variation between the accuracy of the various serological studies included. Including the confidence intervals of each seroprevalence estimate in the comparison allows for some of this variation to be captured quantitatively, but most will be missed. However, as the comparison is crude for a number of reasons, we believe the additional error incurred by such an assumption is minimal. Further, given that our estimates of under-ascertainment in many countries suggest that the numbers of symptomatic infections at the peak of the outbreak were one or two orders of magnitude larger than reported cases, even if deaths are under-reported, our estimates are still likely to be much closer to the true burden than locally reported cases imply.

Our estimates of under-ascertainment over time require a time-series of COVID-19 deaths as an input, a data source that may also exhibit reporting variation. One notable example of this was Spain during June 2020 (Supplementary Appendix: Figure S1). However, as our Gaussian process model quantifies time-varying case ascertainment, it is able to account for positive or negative spikes in reporting (13) (see the Estimating under-ascertainment rates section in the Supplementary Appendix for more details). Finally, our results are limited by the quality of the input data, which is likely to vary in accuracy between countries. However, as we find good agreement between the 95% credible intervals of our estimates and seroprevalence studies, we believe that our model accurately captures some of this variation.

Since the temporal trend in under-ascertainment does not necessarily reflect trends in reported cases or testing effort, evidence synthesis methods such as the one presented here can provide additional insights into whether observed case patterns reflect the underlying epidemic dynamics. In the early stages of outbreaks, this method can provide an indication of whether a large proportion of cases are being detected – and hence whether transmission may be containable with targeted measures such as isolation and contact tracing – or whether transmission is more widespread and a more extensive response is required. Such estimates can also provide insights in the later stages of an outbreak, as they can indicate high levels of detection in countries that have achieved control. For example, in Australia, an adapted version of our model estimated that 80% (95% CrI: 55% – 100%) of cases had likely been ascertained during the outbreak (22). By adjusting for under-ascertainment, it is also possible to reconstruct the temporal dynamics of SARS-CoV-2 internationally. During February and early March 2020, importations of SARS-CoV-2 into the UK came primarily from Italy, Spain and France (28). This is consistent with the inferred progression of infection during this period in our model; we estimated that Italy, Spain, France and Belgium all had over 6.5% of the population infected by 31st March 2020 (28).

## Conclusion

Consistent with other studies (3,20), we estimated that the true numbers of symptomatic cases and infections are appreciably larger than the number of confirmed cases reported (Figures 1 and 2). We also estimated that the timing of the peak level of symptomatic cases may be considerably earlier or later than the raw confirmed case curve suggests (Table 1). Accurate surveillance of an ongoing outbreak is crucial for estimating key epidemiological values such as the reproduction number, and hence evaluating the impact of control measures (21). If reported case numbers do not reflect the shape and magnitude of the underlying epidemic, it may bias estimates of transmission potential and effectiveness of interventions. If levels of under-ascertainment are increasing, early interventions may appear to be more effective than they actually are, which could lead to delays in imposing more stringent measures. Likewise, if ascertainment increases in the declining phase of an epidemic, the effectiveness of interventions may be underestimated, potentially leading to measures remaining in place for longer than they would have been had more accurate data been available.

## Data Availability

The data we use is publicly available online from the European Centre for Disease Control (ECDC) here: https://www.ecdc.europa.eu/en/publications-data/download-todays-data-geographic-distribution-covid-19-cases-worldwide.

## Declarations

### Ethics approval and consent to participate

Not applicable

### Consent for publication

Not applicable

### Availability of data and materials

The data we use is publicly available online from the ECDC (18). The code for the dCFR and under-reporting estimation model can be found here: https://github.com/thimotei/CFR_calculation. The code to read in the under-ascertainment data and to reproduce the figures in this analysis can be found here: https://github.com/thimotei/covid_underreporting.

### Supplementary information

**Additional file 1: Supplementary Appendix 1-4. Figure S1: Figure S1:** Temporal variation in under-reporting for all countries with greater than 10 deaths for more than 50 days. **Figure S2:** Temporal variation in testing effort for all countries there was data for in the Our World In Data database (17). **Figure S3:** the relationship between case ascertainment and testing effort. We define testing effort as the 7-day moving average of the number of new tests per new case each day. We plot the under-ascertainment estimates along with the testing effort estimates for all countries we have both data for. We then fit, using a loess curve to highlight the positive but weak relationship (, where is Kendall’s rank coefficient). **Figure S4:** Temporal variation in under-ascertainment and testing effort for the nine countries with the maximum total cases that we have reliable testing effort estimates for. This figure differs from Figure 1 as the results are computed using the indirectly age-adjusted baseline CFR for each country. **Figure S5:** Confirmed case curves adjusted for temporal under-ascertainment adjusted indirectly for age. The results are similar to those in Figure 2 but have been computed using an indirectly age-adjusted baseline CFR for each country. **Figure S6:** Estimated infection prevalence curves compared with observed seroprevalence data. The results are similar to those in Figure 3 but have been computed using an indirectly age-adjusted baseline CFR for each country. **Figure S7:** Temporal variation in under-reporting for all countries with greater than 10 deaths for more than 50 days. The results are similar to those in Figure S1 but have been computed using an indirectly age-adjusted baseline CFR for each country. **Table S1:** A summary of the country-level serological studies we used for comparison against our model estimates. **Table S2:** A summary of the city-level or regional-level serological studies we used for comparison against our model estimates. **Table S3:** A summary of the parameters, distributions and output quantities either as inputs or outputs of our under-ascertainment model

### Authors’ contributions

TWR and AJK conceived the study. TWK, AJK and NG developed the model and designed the inference framework. TWR wrote the initial draft manuscript with AJK. LW integrated the model into the High Performance Computing (HPC) environment and oversaw all computational aspects of the HPC execution process. All authors read and approved the final manuscript.

## Acknowledgements

The following authors were part of the Centre for Mathematical Modelling of Infectious Disease 2019-nCoV working group. Each contributed in processing, cleaning and interpretation of data, interpreted findings, contributed to the manuscript, and approved the work for publication: Arminder K Deol, C Julian Villabona-Arenas, Thibaut Jombart, Carl A B Pearson, Kathleen O’Reilly, James D Munday, Sophie R Meakin, Rachel Lowe, Amy Gimma, Akira Endo, Emily S Nightingale, Graham Medley, Anna M Foss, Gwenan M Knight, Kiesha Prem, Stéphane Hué, Charlie Diamond, James W Rudge, Katherine E. Atkins, Megan Auzenbergs, Stefan Flasche, Rein M G J Houben, Billy J Quilty, Petra Klepac, Matthew Quaife, Sebastian Funk, Quentin J Leclerc, Jon C Emery, Mark Jit, David Simons, Nikos I Bosse, Simon R Procter, Fiona Yueqian Sun, Samuel Clifford, Katharine Sherratt, Alicia Rosello, Nicholas G. Davies, Oliver Brady, Damien C Tully, Georgia R Gore-Langton.

The authors, on behalf of the Centre for the Mathematical Modelling of Infectious Diseases (CMMID) COVID-19 working group, wish to thank the Defence Science and Technology Laboratory (Dstl) for providing the High Performance Computing facilities and associated expertise that has enabled these models to be prepared, run and processed in an appropriately rapid and highly efficient manner. Dstl is part of the Ministry of Defence.

